# Building a growing genomic data repository for maternal and fetal health through the PING Consortium

**DOI:** 10.1101/2024.05.24.24307899

**Authors:** Clara M. Abdelmalek, Shriya Singh, Blain Fasil, Allison R. Horvath, Sarah B. Mulkey, Carlos Curé, Maribel Campos, Denise P. Cavalcanti, Van T. Tong, Marcela Mercado, Marcela Daza, Mónica Marcela Benavides, Jacqueline Acosta, Suzanne Gilboa, Diana Valencia, Christina L. Sancken, Suzanne Newton, Deolinda M. F. Scalabrin, Marisa M. Mussi-Pinhata, Zilton Vasconcelos, Nahida Chakhtoura, Jack Moye, Elizabeth J. Leslie, Dorothy Bulas, Gilbert Vezina, Fernanda J. P. Marques, Marcio Leyser, Miguel Del Campo, Eric Vilain, Roberta L. DeBiasi, Tongguang Wang, Avindra Nath, Tarik Haydar, Max Muenke, Tamer A. Mansour, Adre J. du Plessis, Jeffrey C. Murray, José F. Cordero, Youssef A. Kousa

**Affiliations:** Center for Genetic Medicine, Children’s National Research Institute, Washington, DC, USA; Center for Neuroscience Research, Children’s National Research Institute, Washington, DC, USA; Honors College, University of Maryland, College Park, MD; Department of Neurology, George Washington University School of Medicine and Health Sciences, Washington, DC, USA; Department of Pediatrics, George Washington University School of Medicine and Health Sciences, Washington, DC, USA; Department of Prenatal Pediatrics Institute, Children’s National Hospital, Washington, DC, USA; BIOMELab, Barranquilla, Colombia; Center for Community Outreach for Health Across the Lifespan, Dental and Craniofacial Genomics Core, Medical Sciences Campus, University of Puerto Rico, San Juan, Puerto Rico; Perinatal Genetics Program, Department of Translational Medicine, Medical Genetics, University of Campinas, Campinas, Brazil; Division of Birth Defects and Infant Disorders, Centers for Disease Control and Prevention, Atlanta, GA, USA; Facultad de Ciencias de la Salud, Universidad del Norte, Barranquilla, Colombia; Universidad Nacional de Colombia, Bogotá, Colombia; Instituto Nacional de Salud, Bogotá, Colombia; Vysnova Partners, Inc., Landover, MD, USA; Division of Preparedness and Emerging Infections, Centers for Disease Control and Prevention, Atlanta, GA, USA; Department of Epidemiology of Microbial Diseases, Yale School of Public Health, New Haven, CT, USA; Gonçalo Moniz Research Center, Oswaldo Cruz Foundation/Ministry of Health, Salvador, Brazil; Department of Pediatrics, Ribeirão Preto Medical School, University of São Paulo, Ribeirão Preto, Brazil; Instituto Fernandes Figueira, Oswaldo Cruz Foundation, Rio de Janeiro, Brazil; Pregnancy and Perinatology Branch, Eunice Kennedy Shriver National Institute of Child Health and Human Development, National Institutes of Health, Bethesda, MD, USA; Maternal and Pediatric Infectious Disease Branch, Eunice Kennedy Shriver National Institute of Child Health and Human Development, National Institutes of Health, Bethesda, MD, USA; Department of Human Genetics, Emory University School of Medicine, Atlanta, GA, USA; Department of Radiology, George Washington University School of Medicine and Health Sciences, Washington, DC, USA; Division of Neuroradiology at Children’s National Hospital, George Washington University School of Medicine and Health Sciences, Washington, DC, USA; SARAH Network of Rehabilitation Hospitals, Rio de Janeiro, Brazil; Stead Family Department of Pediatrics, University of Iowa, Iowa City, IA, USA; Department of Pediatrics, Division of Genetics, University of California, San Diego and Rady Children’s Hospital-San Diego, San Diego, California, USA; Institute for Clinical and Translational Science, University of California, Irvine, CA, USA; Division of Pediatric Infectious Diseases, Children’s National Hospital, Washington, DC, USA; Department of Microbiology, Immunology and Tropical Medicine, George Washington University School of Medicine and Health Sciences, Washington, DC, USA; Translational Neuroscience Center, National Institute of Neurological Disorders and Stroke, National Institutes of Health, Bethesda, MD, USA; Section of Infections of the Nervous System, National Institute of Neurological Disorders and Stroke, National Institutes of Health, Bethesda, MD, USA; National Human Genome Research Institute, National Institutes of Health, Bethesda, MD, USA; Department of Population Health and Reproduction, University of California, Davis, CA, USA; Department of Clinical Pathology, College of Medicine, Mansoura University, Egypt; Department of Epidemiology and Biostatistics, College of Public Health, University of Georgia, Athens, GA, USA; Division of Neurology, and Critical Care, Children’s National Hospital, Washington, DC, USA; Neonatal Neurology Program, and Critical Care, Children’s National Hospital, Washington, DC, USA; Division of Neurophysiology, Epilepsy, and Critical Care, Children’s National Hospital, Washington, DC, USA; Department of Genomics and Precision Medicine, George Washington University School of Medicine and Health Sciences, Washington, DC, USA

**Author notes:** **Correspondence to:** Youssef A. Kousa, MS, DO, PhD, Center for Genetic Medicine, Children’s National Research and Innovation Campus, 7144 13^th^ Pl NW, Suite 1247, Washington, DC 20012. **CDC Disclaimer:** The findings and conclusions in this report are those of the authors and do not necessarily represent the official position of the Centers for Disease Control and Prevention.

**Keywords:** virally induced prenatal brain injury, developmental disabilities, brain maturation, Zika

## Abstract

**Background:** Prenatally transmitted viruses can cause severe damage to the developing brain. There is unexplained variability in prenatal brain injury and postnatal neurodevelopmental outcomes, suggesting disease modifiers. Discordant outcomes among dizygotic twins could be explained by genetic susceptibly or protection. Among several well-recognized threats to the developing brain, Zika is a mosquito-borne, positive-stranded RNA virus that was originally isolated in Uganda and spread to cause epidemics in Africa, Asia, and the Americas. In the Americas, the virus caused congenital Zika syndrome and a multitude of neurodevelopmental disorders. As of now, there is no preventative treatment or cure for the adverse outcomes caused by prenatal Zika infection. The Prenatal Infection and Neurodevelopmental Genetics (PING) Consortium was initiated in 2016 to identify factors modulating prenatal brain injury and postnatal neurodevelopmental outcomes for Zika and other prenatal viral infections.

**Methods:** The Consortium has pooled information from eight multi-site studies conducted at 23 research centers in six countries to build a growing clinical and genomic data repository. This repository is being mined to search for modifiers of virally induced brain injury and developmental outcomes. Multilateral partnerships include commitments with Children’s National Hospital (USA), *Instituto Nacional de Salud* (Colombia), the Natural History of Zika Virus Infection in Gestation program (Brazil), and Zika *Instituto Fernandes Figueira* (Brazil), in addition to the Centers for Disease Control and Prevention and the National Institutes of Health.

**Discussion:** Our goal in bringing together these sets of patient data was to test the hypothesis that personal and populational genetic differences affect the severity of brain injury after a prenatal viral infection and modify neurodevelopmental outcomes. We have enrolled 4,102 mothers and 3,877 infants with 3,063 biological samples and clinical data covering over 80 phenotypic fields and 5,000 variables. There were several notable challenges in bringing together cohorts enrolled in different studies, including variability in the timepoints evaluated and the collected clinical data and biospecimens. Thus far, we have performed whole exome sequencing on 1,226 participants. Here, we present the Consortium’s formation and the overarching study design. We began our investigation with prenatal Zika infection with the goal of applying this knowledge to other prenatal infections and exposures that can affect brain development.

## INTRODUCTION

### Prenatal Viral Infection

Prenatal viral infections are a leading cause of preterm birth, growth restriction, fetal demise, and spontaneous abortion.^1-4^ In developing countries, upwards of one in every four pregnancies is exposed to viral infections.^5,6^ Prenatal infections are of utmost concern because they are difficult to diagnose and the developing brain is uniquely susceptible to injury. When the prenatal brain is injured, a host of serious neurodevelopmental disabilities can ensue, which are life-altering. Although the full range of clinical outcomes are still being determined, we know that some infants experience moderate to severe neurodevelopmental disabilities, while others appear to be unaffected at birth.^7,8^ Variability in brain injury and neurodevelopmental delay after a prenatal infection suggests modifiers. Identification of these modifiers can inform the development of therapies that can be implemented before or after birth. Despite the epidemiological and clinical implications of the problem, there are few prenatal viral infections for which there is any treatment. For example, cytomegalovirus (CMV) antivirals are used for symptomatic children to reduce severity of hearing loss.^9^ Consequently, there is an urgent need for therapies that both prevent and treat prenatal viral infections.

Variability of clinical outcomes is likely influenced by multiple factors at both the individual and populational levels. Individual factors include the specific virus, viral load, and gestational age at time of infection. Populational factors include the public health infrastructure and community, and others that exist among a complex interplay. Interactions between individual and populational factors are likely.^7,10,11^ Today, the contributions of host genetic modifiers are unknown.^12^ However, the information is critically needed because it can inform our basic and mechanistic understanding of prenatal viral infections and aid in identifying patients at highest risk of having an affected infant(s).^13-15^ Combined, these insights may pave a road toward development and delivery of much needed therapies.

### Zika Virus

Among a growing number of recognizable viral infections that can affect brain development, Zika virus emerged as an international public health emergency after serial epidemics were first detected in 2013. After spreading through Africa, Asia, and the Americas, the virus is now endemic in many countries in the Southern Hemisphere.^8^ During a Brazilian epidemic, prenatal Zika virus infection was found to be associated with severe brain injury leading to microcephaly, a condition defined by an infant’s head circumference measuring two/three standard deviations below the mean for gestational age, sex, and ethnicity.^10^ Novel viral mutations increased virulence and transmissibility, which likely increased pathogenicity during the epidemic.^16^ Prenatal Zika infection was later identified as the cause of congenital Zika syndrome, a clinical condition associated with severe microcephaly, a partially collapsed skull, and limb, ocular, and hearing abnormalities.^17^ As typical of other prenatal viral infections, these outcomes can occur in isolation or as a combination of phenotypes. Importantly, brain malformations are associated with lifelong morbidity and an increased risk of mortality. By investigating Zika, we aim to expand upon our current knowledge of virally induced neuronal injury and the developing brain.

### The Case for Genetic Susceptibility

In the case of Zika virus, modifiers of brain injury and neurodevelopmental outcome are unknown.^18-27^ During the 2015 epidemic, outcomes of prenatally infected monozygotic twins were more likely to be concordant compared to those of dizygotic twins, consistent with genetic differences in the twins contributing to susceptibility.^22^ Further, outcomes of related individuals were more likely to be concordant compared to those of unrelated individuals.^21,28-30^ Therefore, differences in gene expression or genetic regulation in response to viral infections might contribute to differences in neurodevelopmental outcomes. As with other infections, identifying susceptibilities informs our understanding of viral replication, cell injury, and ultimately, disease severity. Thus, inherited susceptibility might be an important determinant of neurodevelopmental attainment.

### Complexities in Testing the Hypothesis

Our understanding of genetic susceptibility in the prenatal period remains incomplete because of the multifaceted dynamics inherent to maternal and prenatal health. There are also cost and statistical limitations in testing for a genetic association with postnatal neurodevelopmental delays because a sufficiently powered study requires both a large participant cohort and long-term clinical follow-up.^31-34^

### Prenatal Infection and Neurodevelopmental Genetics (PING) Consortium

To address an inherently complex research gap, we formed a team composed of conceptual and technical subject matter experts covering topics ranging from viral neurobiology and prenatal neurology to populational and global health and genomics. We established the Prenatal Infection and Neurodevelopmental Genetics Consortium (PING) in 2016 (formerly Zika Genetics Consortium) with a group of international experts in these diverse fields. The original mission of the Consortium was to identify maternal/fetal risk or protective factors in brain injury caused by Zika. Moving forward, the team has also decided to leverage the current framework to address critical questions toward preventing prenatal viral infections, brain injury, and neurodevelopmental disorders.

A genomic association study requires a large participant cohort. To build the cohort, the Consortium partnered with international studies through bilateral and multilateral commitments. Bilateral commitments include sites in Colombia, Puerto Rico, and Brazil through collaboration with Children’s National Hospital in Washington, DC. Multilateral commitments include the *Vigilancia de Embarazadas con Zika* (VEZ) and *Zika en Embarazadas y Niños* (ZEN) studies, launched by Colombia’s *Instituto Nacional de Salud* (INS) and the U.S. Centers for Disease Control and Prevention (CDC); the Natural History of Zika Virus Infection in Gestation (NATZIG) cohort study launched by the *Universidade de São Paulo*, Brazil; the Zika *Instituto Fernandes Figueira* cohort launched by Fiocruz in Rio de Janeiro; and the Zika in Infants and Pregnancy (ZIP) study launched by the U.S. National Institutes of Health (NIH). Today, the Consortium represents six countries and 22 research cohort sites (Figure 1) and the biorepository contains samples from diverse and understudied patient cohorts representative of urban, suburban, and rural South and Central American populations.

**Figure 1.**
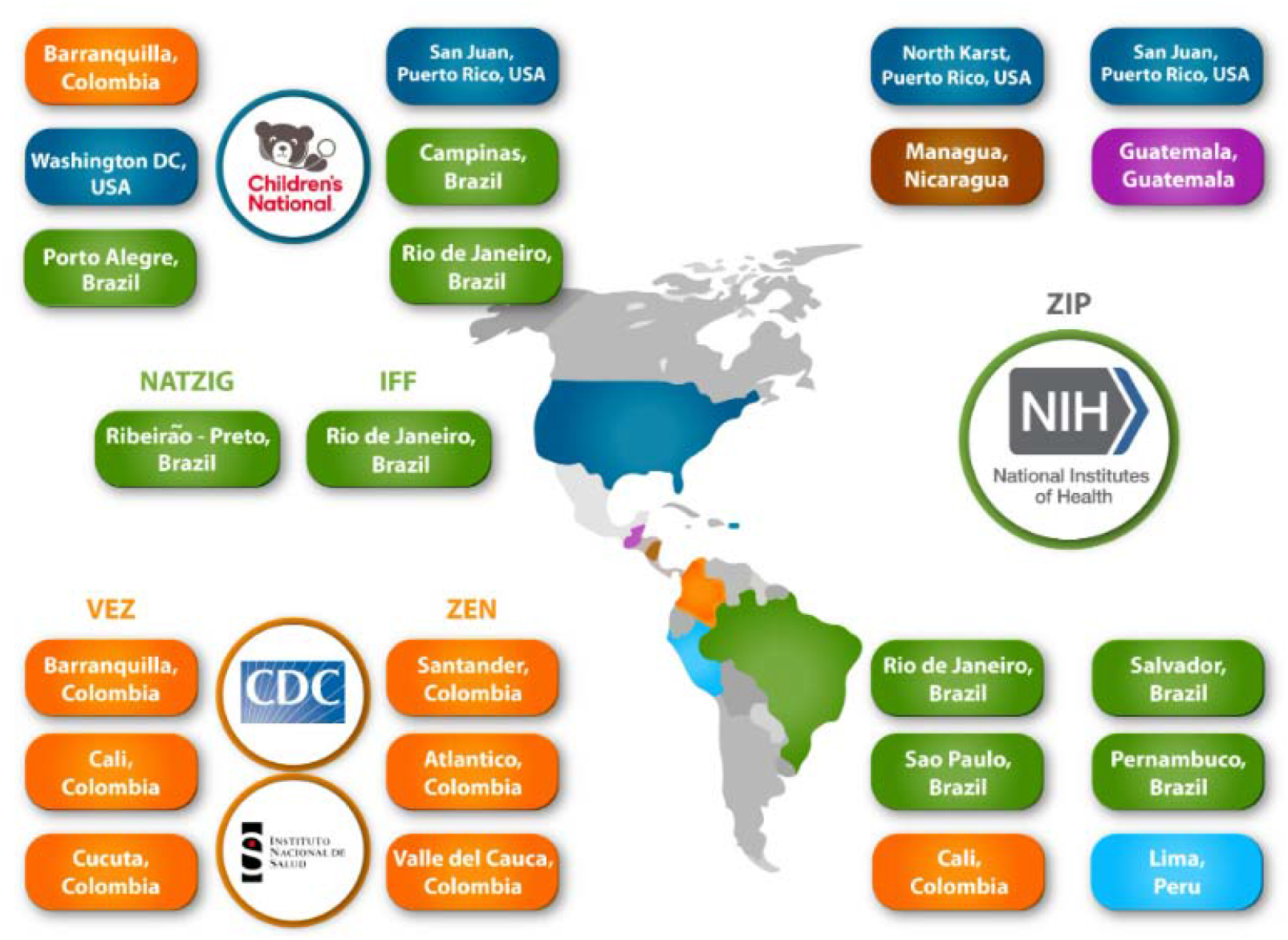
Partnerships and Sites. Through collaborations at Children’s National Hospital with the U.S. Centers for Disease Control and Prevention, Colombia *Instituto Nacional de Salud*, the Natural History of Zika Virus Infection in Gestation cohort, Zika *Instituto Fernandes Figueira cohort*, and the National Institutes of Health, the Consortium represents six countries and 22 research cohort sites (there are two ZIP cohorts in Puerto Rico). For the purposes of the Consortium, Puerto Rico is considered as a US territory.

Our short-term goal is to identify through our established protocol host genetic risk and protective factors modifying virally induced prenatal brain injury. This objective can provide several longer-term benefits, including: 1) improving our understanding of brain development in the context of prenatal viral infections; 2) clarifying the molecular pathway of infection by which a virus evades immune and cellular response; 3) elucidating the mechanism of cellular and neuronal injury; 4) connecting the pathophysiology of prenatal brain injury with postnatal neurodevelopmental outcomes; 5) establishing more precision in predicting developmental delays; and 6) aiding in designing targeted therapies for specific viral infections and individuals at greatest risk. Ultimately, a deeper understanding of host modifiers is crucial for the development of targeted therapies to prevent brain injury.

### Broader Applicability

Broader applicability is expected from our investigation of prenatal Zika infection for several reasons. First, many prenatal viral infections often result in overlapping neuropathology and clinical outcomes.^35^ Thus, the proposed work will deepen our understanding of other prenatal viral infections. Second, Zika is associated with a severe range of neurodevelopmental disorders, making it an exemplar for studying if and how viruses can interact with prenatal neurogenetic programming to affect brain development.^17^ Third, as ongoing work evaluates the impact of SARS-CoV-2 on human development, we are still not adequately prepared to protect the developing brain in future pandemics. Further, even for known prenatal viral infections with available vaccines (e.g. rubella), declining vaccinations rates could lead to pathogen re-emergence, which could have devastating effects on the prenatal brain similar to Zika virus. Finally, as the Consortium develops further with sites throughout the western hemisphere, there might be opportunities for early surveillance and monitoring for emerging or remerging infection affecting prenatal development. The work started here may advance our understanding of viral neurobiology, which is essential in designing neuroprotective strategies and antiviral therapies.

## PROTOCOL METHODS

### Overarching Study Design

For our present work, the Consortium designed a case-control study to identify causes of variability in and modifiers of virally induced prenatal brain injury and postnatal neurodevelopmental outcomes. However, there were several challenges to consider. First, serial epidemics were affecting multiple populations in different countries over the course of several years, with varying rates of exposure, infection, and outcome. These differences amplified the need for diversity in the participant cohort. Second, rates of infection declined in 2017, and most infected individuals were born before prospective studies were approved and started enrollment. Third, as mentioned above, genomic studies require large participant cohorts to generate enough statistical power during data analysis.

Each of these considerations were addressed in the study design. First, toward a more diverse and representative participant cohort, we partnered with multiple research centers throughout Central and South America. To account for the declining rates of infection, our study included both prospective studies, which primarily enrolled unexposed individuals, and retrospective studies, which enrolled exposed and affected individuals. In addressing the first two points, our participant cohort needed to become larger. Consequently, we met a third objective, as a larger participant cohort provided more statistical power for genomic data analysis. In several cases, studies extended each other synergistically. For instance, while some cohorts had prenatal imaging, others performed longer-term and more extensive follow-up on affected and/or unaffected infants. Linking the two, we had more information on prenatal correlates of neurodevelopmental outcomes.

The Consortium gained access to clinical data and biospecimens from several existing studies (Table 1), reducing startup costs and extending the benefit of intricately designed parent studies. Clinical data included pre- and postnatal neuroimaging, subclinical/morphometric phenotyping, and up to two years of neurodevelopmental follow-up. Biospecimens included blood, urine, and placenta samples, which were banked for ongoing research purposes. Whole exome sequencing was performed on collected samples, with the goal of creating a databank that will serve as a resource in current and future genetic research on prenatal viral infections (Figure 2). To account for the inherent limitations of whole exome sequencing, the Consortium will pursue additional types of sequencing in the future.

**Table 1.** Maternal and Infant Assessments.

| **Assessment** | **Data Collected (as available)** |
| --- | --- |
| **Maternal Data** |  |
| Pregnancy and medical history | Date of birth, gender, race, ethnicity, number of prior pregnancies and their outcomes, time since last pregnancy and medical/mental health history, including chronic and gestational clinical problems and current/prior medications |
| Laboratory testing | Serological examination and/or RT-PCR, for Zika and other prenatal infectious conditions |
| Biological samples | Any sample for DNA extraction, collected at any point during the study period |
| **Infant Data** |  |
| Birth and demographic history | Date of birth, gender, race, ethnicity, gestational age at birth, type of delivery, complications in childbirth or in the neonatal period |
| Clinical and developmental data | Family history, medications, other co-occurring medical conditions, growth parameters (weight, length, and head circumference), and neurodevelopmental assessment (measured by any validated neurodevelopmental tool) obtained sequentially or at a single time point |
| Hearing and ophthalmological assessments | Auditory or auditory brainstem response and general eye examinations |
| Neuroimaging examinations | Prenatal or postnatal ultrasounds, CT scans, and MR imaging |
| Biological samples | Any sample for DNA extraction, collected at any point during the study period |

**Figure 2.**
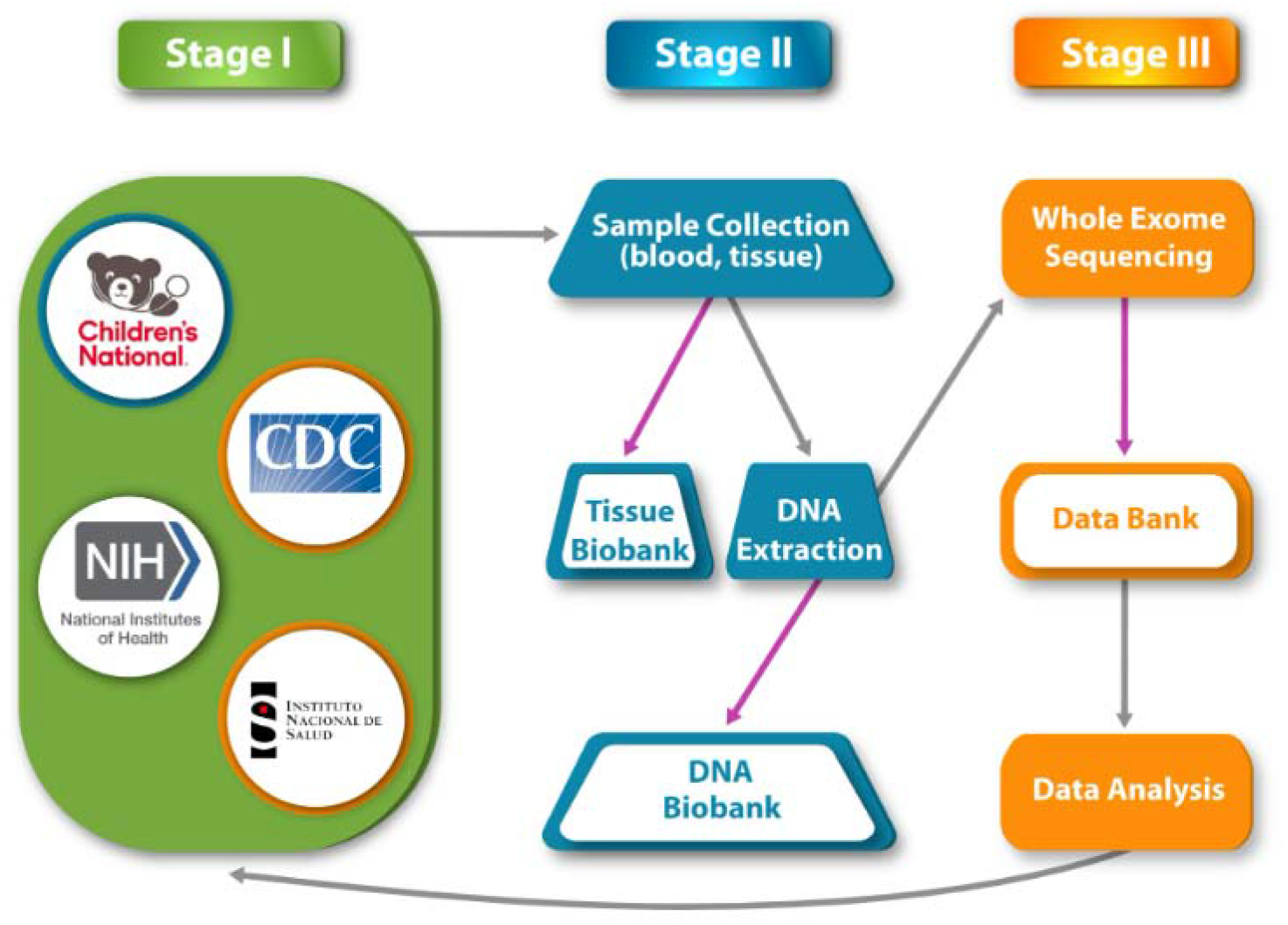
Data Collection, Processing, and Storage. As listed in Stage I, the Consortium gained access to clinical data and biospecimens. Clinical data available among the studies included pre- and postnatal imaging and subclinical phenotyping, and up to two years of neurodevelopmental follow-up. As described in Stage II, biospecimens, which included blood, tissue, and placenta samples, were collected for DNA extraction. Tissue and DNA biobanks were established to preserve samples for future research purposes. Whole exome sequencing was performed on collected samples, with the goal of creating a databank that will serve as a resource in current and future research on prenatal viral infections, as represented in Stage III. Grey arrows refer to processes, purple arrows indicate biobank storage.

### Studies Pooled Mother-Infant Populations

We grouped mother-infant dyads from the feeding studies according to both viral infection status and phenotypic outcome as follows: 1) unexposed mother/unaffected child; 2) exposed mother/unaffected child; and 3) exposed mother/affected child. We obtained access to unexposed dyads from prospective studies, and these participants served as a reference population for our case-control study. Retrospective cohort studies provided access to exposed dyads. Exposed-unaffected individuals were represented in both study types. As a function of pair-wise comparisons among the three groups, we will evaluate for both maternal and/or prenatal genetic modifiers––in line with the established protocol. When available, we included unexposed mother/affected infant dyads as a reference population to identify causal genetic variants for syndromic disorders that could include microcephaly without an associated viral etiology.

### Statistical Consideration

We conducted a power analysis to determine the sample size needed for association testing between clinical outcome and rare/common variants in candidate genes. These considerations informed our study design. For instance, to detect a genetic association with alleles having an odds ratio >1.5 and minor allele frequencies >5% with 80% power, a sample size of 1,000 individuals are needed (500 cases, 500 controls).

### Defining Exposure

As commonly observed, laboratory assays for Zika virus evolved during the epidemic. Even today, the specificity and sensitivity of testing for prenatal Zika infection is not optimized, with molecular testing (PCR) requiring more proximity to time of infection, and serological testing (antibody) having high cross-reactivity with related flavivirus infections (Dengue, in particular) and assays (e.g., PRNT). As a such, it was difficult to clearly delineate the timing and type of exposure in some cases. For these reasons, we define exposure as a pregnant woman who tested positive for Zika by PCR or antibody testing in the correct sensitivity window and/or had symptoms consistent with Zika infection during the epidemic. We define unexposed as a pregnant woman who tested negative for Zika by PCR or antibody testing and had no history of Zika virus infection or similar symptoms during pregnancy. A clinical diagnosis of Zika virus infection was used in determining cases and controls. As availability of collected data permits, we are evaluating how co-infection of Dengue, chikungunya, and SARS-CoV-2 modified prenatal brain development and neurological outcomes.

Some limitations are noted. First, testing was limited early in the epidemics and in certain geographic areas. In addition, both molecular and serological assays had limited sensitivity and specificity for Zika. *Fourteen sites included testing for other known infections, confirming that outcomes are likely caused by Zika infection. If sites did not have additional testing, we will perform a multi-variate linear regression analysis to address the potential confounding of co-occurring infections*. Finally, as generally agreed, at least 80% of pregnant women were asymptomatic for infection.^36^ These limitations lead to under-detection of exposed pregnant women and infants.

### Defining Outcomes

Prenatal Zika infection can result in a spectrum of phenotypes.^7,8^ We considered a participating infant affected if they had clinical or neurodevelopmental testing consistent with developmental delay, learning or intellectual disability or imaging characteristics of a prenatal viral infection, such as intracranial calcifications, ventriculomegaly, cerebral atrophy, cortical malformation, vasculopathy, or microcephaly. Infants were considered unaffected if they had typical development, normal cognitive function, and/or normal imaging (if available).

### Inclusion/Exclusion Criteria

We included all eligible participants regardless of religion, gender, or ethnic origin. Primary inclusion and exclusion criteria were set by parent/partnering studies, as previously described.^7,38,39^ We included infants enrolled and referred to our study as long as they met the outcome criteria. In the primary analyses, we excluded mother-infant dyads if the pregnant woman did not have a history of exposure and her infant was affected by a disorder likely unrelated to prenatal Zika infection. Previously collected and deidentified clinical data was used to participants.

### Sample Quality Control

Participation in this study is limited to individuals who provide a biospecimen for future studies. Once a biospecimen is identified, DNA extraction is performed on a portion of the provided sample. DNA integrity, quality, and purity is evaluated using standard lab techniques including Qubit Assay, DNA TapeStation and Nanodrop. DNA quality is evaluated with spectrophotometry, with an ideal absorbance rate of 1.8-2.0 at 260/280 nm. Samples that pass quality control testing are sequenced through existing protocols in the PING Consortium at Children’s National Hospital in Washington, DC.1Biospecimens used for genetic analysis are stored locally at the primary study site or with the Consortium’s biorepository in Washington, DC. The samples are kept at a -80°C freezer, protected with an access key. Samples are deidentified and only the primary-based research team in each site has access to the data that links patient identifiers with the de-identified information.

### Whole Exome Sequencing Pipeline

Approved samples will be used for preparation of sequencing libraries using Agilent SureSelect Human All ExonV6 kit (Agilent Technologies, CA, USA) following manufacturer’s recommendations. The library will be checked with Qubit and real-time PCR for quantification and Agilent Bioanalyzer for size distribution detection. Qualified libraries will be sequenced at Novogene (Sacramento, CA) on Illumina platforms with PE150 strategy. Germline variant calling will be done using the DRAGEN-GATK best practice workflow of the Genome Analysis Toolkit (GATK v4.1.9.0).^40^

### Planned Genetic Association Study Design

To evaluate for maternal and fetal single nucleotide polymorphisms that contribute to risk and protection, we will do at least two types of genomic analyses. First, we will perform a case/control genetic association study. Here, we will compare dyads that are exposed/affected to those that are exposed/unaffected. Next, we will perform an association study to identify modifiers of quantitative traits (e.g., head circumference). Here, we will compare unexposed and exposed individuals with mild, moderate, and severe outcomes among each other. Continuing with the above example, we will include known and possible variables in the outcome to determine genetic factors that contribute to both a smaller head circumference and the severity of microcephaly as clinically defined. Both types of analyses can be done within individual cohorts and across the Consortium’s cohort. We will evaluate for ancestral differences by correcting for population stratification. As far as power analyses permit, we will test for associations with both rare and common alleles.

To account for the wide spectrum of phenotypic outcomes after infection, we will compare unaffected and affected individuals in parallel experimental design. In doing so, we will determine susceptibility to any negative outcome (isolated/combinatorial, mild/severe), as well as to specific outcomes. For example, we will compare exposed/unaffected individuals to all those who are exposed/affected and compare exposed/unaffected individuals to those with specific phenotypes, including microcephaly or ventriculomegaly.

## THE CONSORTIUM’S RESEARCH PARTNERSHIP

The PING Consortium established a protocol for data and sample gathering from the 23 research centers, coordinated by a sub-group of partnering entities. What follows is a description of patient-cohort sites from which data have been gathered, along with their liaising entities.

### Barranquilla, Colombia – Children’s National Hospital

#### Overview

Beginning in 2016, Dr. Sarah Mulkey and colleagues began a prospective cohort study to investigate prenatal and postnatal neuroimaging and longitudinal neurodevelopmental outcomes in children with prenatal Zika virus exposure. The study included pregnant participants with Zika virus infection in Atlántico Department, Colombia, and in Washington, D.C. (émigrés to USA or returning travelers). The pregnant mother-infant dyads were enrolled and followed through pregnancy and delivery and had pre- and postnatal neuroimaging and neurodevelopmental assessments (Table 2). Infants with antenatal Zika virus exposure, but without congenital Zika syndrome were then followed longitudinally to assess long-term neurodevelopmental outcomes in both the U.S. and in Colombia. Non-Zika virus exposed children were enrolled in the U.S. and in Colombia as controls at age four years and will undergo similar longitudinal assessments for comparison to the Zika-exposed cases up to age seven years.

**Table 2.**
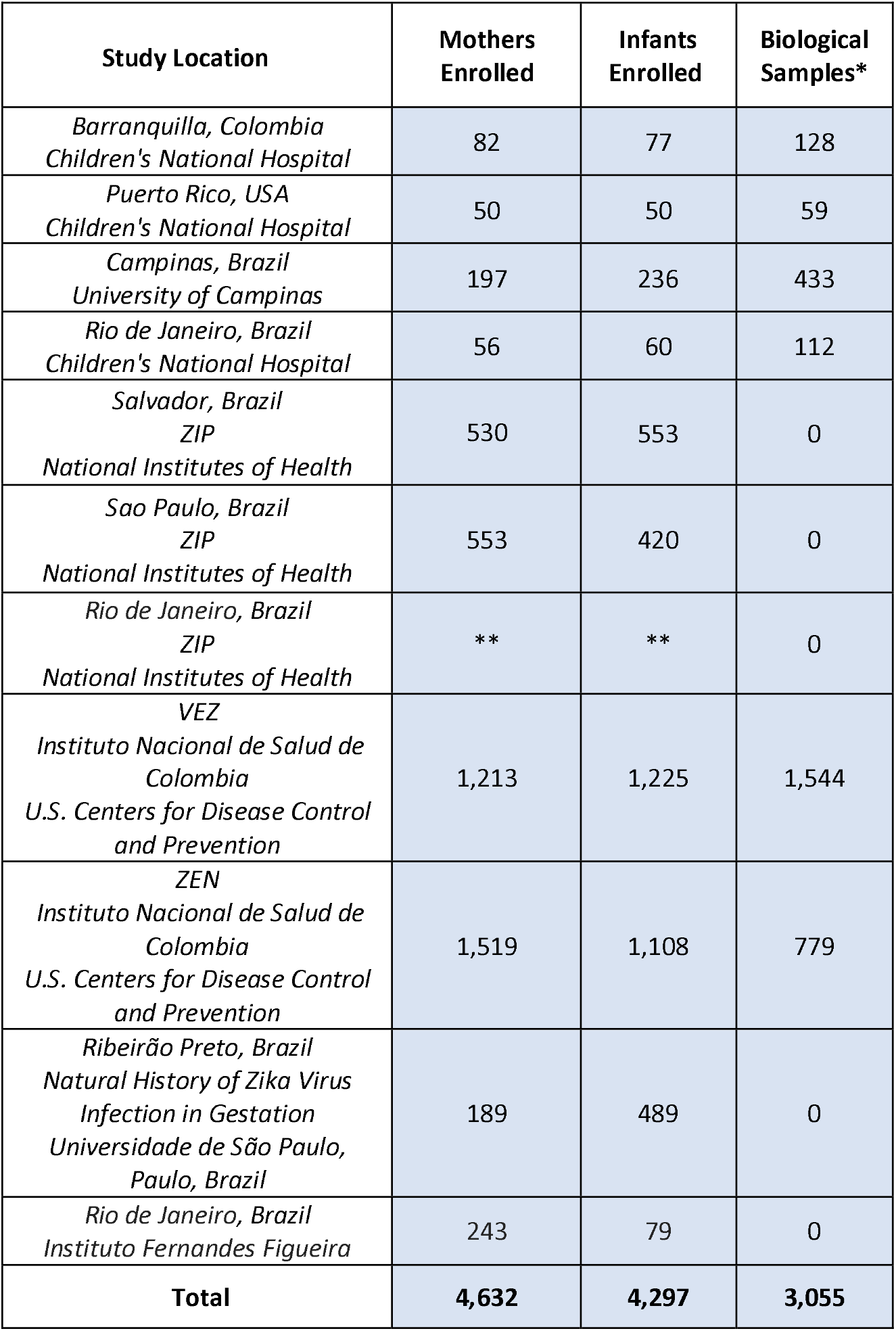
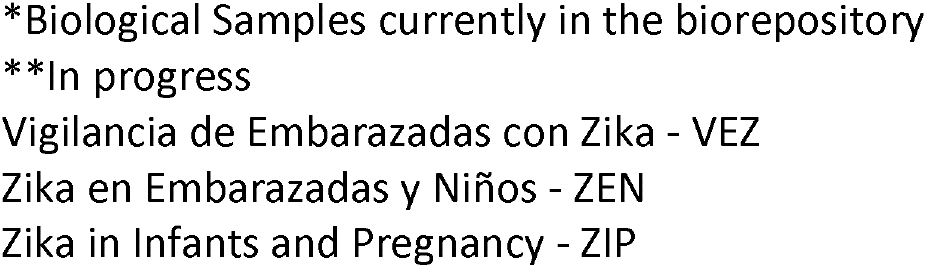
Participant Enrollment Numbers and Participants Sampled.

| **Study Location** | **Mothers Enrolled** | **Infants Enrolled** | **Biological Samples\*** |
| --- | --- | --- | --- |
| *Barranquilla, Colombia*<br>*Children's National Hospital* | 82 | 77 | 128 |
| *Puerto Rico, USA*<br>*Children's National Hospital* | 50 | 50 | 59 |
| *Campinas, Brazil*<br>*University of Campinas* | 197 | 236 | 433 |
| *Rio de Janeiro, Brazil*<br>*Children's National Hospital* | 56 | 60 | 112 |
| *Salvador, Brazil*<br>*ZIP*<br>*National Institutes of Health* | 530 | 553 | 0 |
| *Sao Paulo, Brazil*<br>*ZIP*<br>*National Institutes of Health* | 553 | 420 | 0 |
| *Rio de Janeiro, Brazil*<br>*ZIP*<br>*National Institutes of Health* | ** | ** | 0 |
| *VEZ*<br>*Instituto Nacional de Salud de*<br>*Colombia*<br>*U.S. Centers for Disease Control*<br>*and Prevention* | 1,213 | 1,225 | 1,544 |
| *ZEN*<br>*Instituto Nacional de Salud de*<br>*Colombia*<br>*U.S. Centers for Disease Control*<br>*and Prevention* | 1,519 | 1,108 | 779 |
| *Ribeirão Preto, Brazil*<br>*Natural History of Zika Virus*<br>*Infection in Gestation*<br>*Universidade de São Paulo,*<br>*Paulo, Brazil* | 189 | 489 | 0 |
| *Rio de Janeiro, Brazil*<br>*Instituto Fernandes Figueira* | 243 | 79 | 0 |
| **Total** | **4,632** | **4,297** | **3,055** |

\*Biological Samples currently in the biorepository
\*\*In progress
Vigilancia de Embarazadas con Zika - VEZ
Zika en Embarazadas y Niños - ZEN
Zika in Infants and Pregnancy - ZIP

Of the Consortium’s 22 committed sites, we have included the 11 sites that have submitted IRBs for a genomic association study in this table.

#### Maternal Follow-Up

Placental tissue and blood samples were collected. All women had confirmatory evidence of Zika virus infection via PCR, IgM, IgG, and/or plaque-reduction neutralization assay.

#### Infant Follow-Up

Blood samples were obtained from prenatally exposed infants, and head circumference and weight were measured serially throughout the study. Neurodevelopmental attainment up to age 18 months was assessed by the Warner Initial Developmental Evaluation of Adaptive and Functional Skills (WIDEA) and the Alberta Infant Motor Scale (AIMS). The Pediatric Evaluation of Disability Inventory Computer Adaptive Test (PEDI-CAT) and Movemenet Assesement Battery for Children (MABC) were used to evaluate mobility of cases and controls between the ages 3-5. Previous MRIs and cranial ultrasonography scans were abstracted for the study. As the controls in this study were not enrolled until the age of 4, there was no imaging available for this group. No biological samples or prenatal imaging is available for the control group.^7, 41^

#### Key Attributes

This cohort has several key features in the clinical data collected. Among them, infants underwent prenatal/neonatal neuroimaging and multi-domain long-term neurodevelopmental follow-up. Limitations include that not all participating children had postnatal imaging, and placentas were not collected from all infants. However, serial follow-up of this cohort continued into preschool age and continues into school-age.

### Puerto Rico, USA – Children’s National Hospital

#### Overview

In 2017, Dr. Maribel Campos and her team designed a prospective case-control study to characterize the impact of Zika infection on prenatal craniofacial and dental development. Pregnant women with Zika exposure receiving care at sites collaborating with the University of Puerto Rico were enrolled and followed through pregnancy and delivery. Exposed cases and unexposed controls were matched by maternal age group, health and socioeconomic status, and history of toxic exposures. Infants were enrolled if their mothers were willing to have a complete postpartum oral health assessment. A total of 50 prenatally exposed infants (25 cases and 25 controls) were enrolled and followed for 12 months postpartum and clinical outcomes were evaluated (Table 2).

#### Maternal Follow-Up

Maternal data describing oral health examinations were collected. In addition, whole blood and saliva samples were obtained (Table 3).

**Table 3.** Maternal Study Design Protocol.

| Maternal Variables |  |  |  |  |  |  |  |
| --- | --- | --- | --- | --- | --- | --- | --- |
|  | Data Collection Method | Enrollment | Trimester |  |  | Delivery | 6 weeks post-partum |
|  |  |  | 1<sup>st</sup> | 2<sup>nd</sup> | 3<sup>rd</sup> |  |  |
| Pregnancy & Medical History | Survey & Medical Records | ◇ □ ○ △ ☆ | ◇ ○ ☆ | ◇ ○ ☆ | ◇ ○ ☆ | ◇ ☆ | ◇ ☆ |
| Laboratory Testing | PCR | ◇ □ ○ ☆ | ◇ ○ ☆ | ◇ ○ ☆ | ◇ ○ ☆ | ◇ ○ ☆ | ◇ ☆ |
|  | Serology | ◇ □ ☆ | ◇ ☆ | ◇ ☆ | ◇ ☆ | ◇ ☆ | ◇ ☆ |
| Biological Samples | Amniotic Fluid, Breast Milk, Blood, Placenta, Saliva, Serum, tissue, Vaginal Swab or Urine | ◇ □ ○ △ ☆ | ◇ □ ○ ☆ | ◇ □ ○ ☆ | ◇ □ ○ ☆ | ◇ □ ○ △ ☆ | ◇ ☆ |

**Key:** □ ZIP; □ VEZ; ○ ZEN; □ CNH – PR; □ NATZIG

#### Infant Follow-Up

Upon enrollment, infant medical records, prenatal/delivery care records, and newborn assessments were abstracted for the study. Oromotor functional assessments were completed at six and 12 months of age. Data from oral health examinations and samples of whole blood and saliva were collected (Table 4).

**Table 4.**
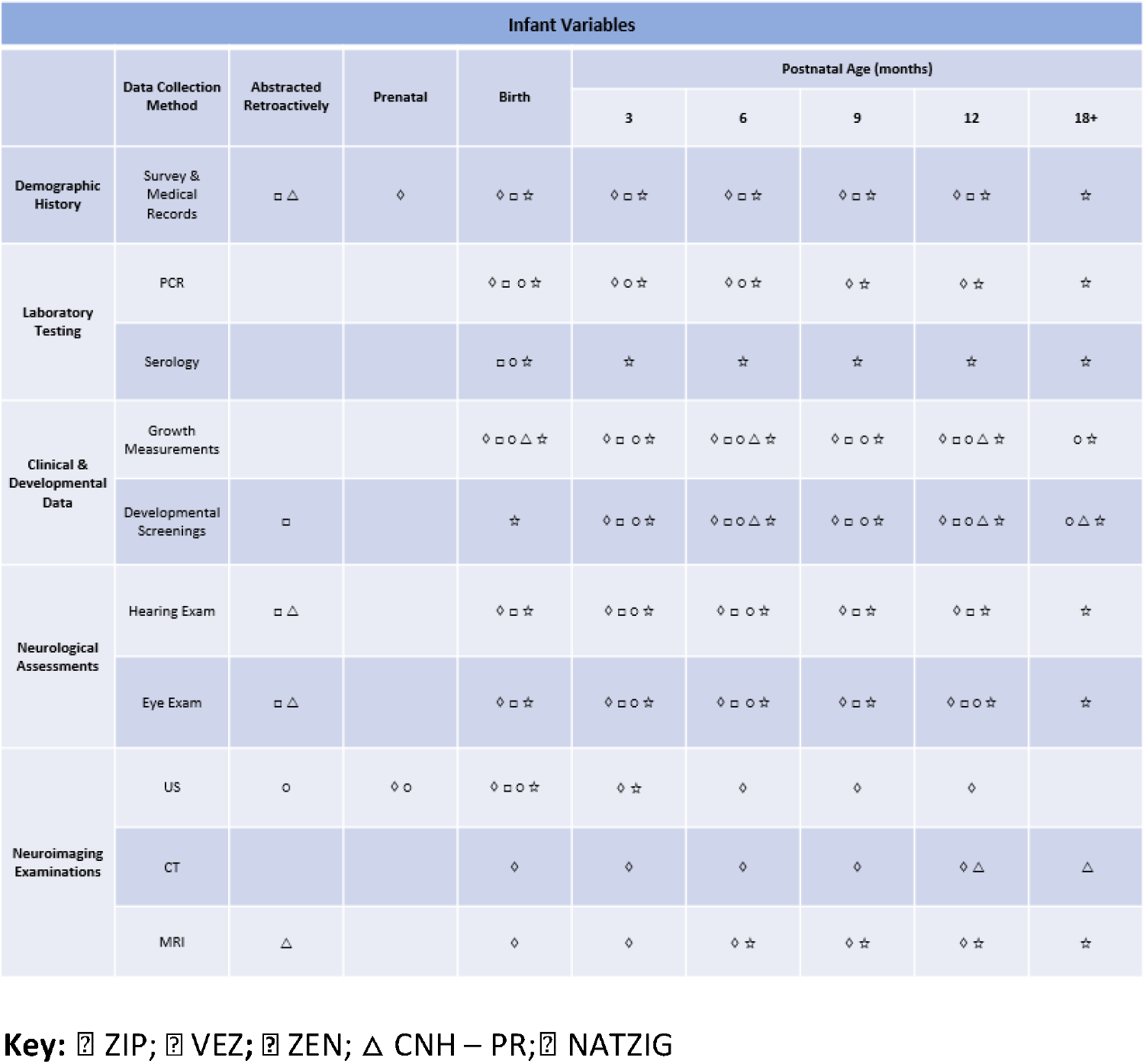
Infant Study Design Protocol.

#### Key Attributes

*The study investigated oral health and craniofacial development in children with congenital Zika syndrome. Neurodevelopmental outcomes were not a focus of this study*.

### Campinas, Brazil – Children’s National Hospital

#### Overview

*Dr. Denise Cavalcanti and her team at the University of Campinas started a retrospective cohort study in 2017 to investigate susceptibility to congenital Zika syndrome after prenatal infection. In addition, the project also aimed to investigate the metabolomics of exposed children and the microbiome of their mothers. The team enrolled infants born to women who exhibited Zika-like symptoms during pregnancy during the 2015-2016 outbreak from three Brazilian cities (Fortaleza, São Luís, João Pessoa). The study enrolled 197 mothers and 236 infants (Table 2). Of these, 208 children had severe neurodevelopmental delays, while 28 children did not present with any delay. Metabolomic data allowed the researchers to identify three biomarkers that suggest that this population suffered from an important inflammatory process; with the detection of mediators associated with glial activation, they proposed that microcephaly is a product of immune response to the virus, as well as excitotoxicity mechanisms, which remain ongoing even after birth*.^42^

#### Maternal Follow-Up

Samples of blood and saliva were collected between May 2017 and January 2020. Some women had confirmatory evidence of Zika infection via laboratory testing (PCR and/or IgM) within the same timeframe the biological samples were collected.

#### Infant Follow-Up

Blood and saliva samples were collected between May 2017 and January 2020. Most children underwent neuroimaging.

#### Key Attributes

The Brazil cohort enrolled a large population of clinically diagnosed, severely affected children. However, some children did not have imaging or neurodevelopmental follow-up.

### Rio de Janeiro – Children’s National Hospital

#### Overview

To investigate functional impairments in children diagnosed with cerebral palsy as a result of congenital Zika syndrome, Drs. Marques and Leyser designed a comprehensive longitudinal study to evaluate clinical and functional manifestations of the disease. Beginning in 2015, the study retrospectively enrolled 56 mothers and 60 children with cerebral palsy after prenatal Zika virus infection (Table 2). Affected children are followed and managed at a tertiary rehabilitation center in Brazil (SARAH).

#### Maternal Follow-Up

Blood samples were collected and Zika-like symptoms during pregnancy were obtained.

#### Infant Follow-Up

Blood samples from infants were collected. To evaluate brain development and for injury, children underwent neuroimaging. Neurodevelopmental assessments included testing for gross motor function (GMFCS), manual ability (MACS), communication (CFCS), eating and drinking ability (EDACS), and visual function (VFCS).

#### Key Attributes

The study enrolled children clinically diagnosed with congenital Zika syndrome and cerebral palsy. As a result, most children were severely affected. Neuroimaging was obtained and the children had neurodevelopmental follow-up.

### Vigilancia de Embarazadas con Zika – Instituto Nacional de Salud de Colombia, U.S. Centers for Disease Control and Prevention

#### Overview

The *Vigilancia de Embarazadas con Zika* project was an intensified surveillance of 1,213 pregnant women with symptoms of prenatal Zika infection in Colombia. The study is aimed at evaluating the relationship between symptoms during pregnancy and adverse pregnancy, birth, and infant outcomes, including birth defects and neurodevelopmental disabilities. Initiated in 2016, VEZ was a collaboration between the Colombian *Instituto Nacional de Salud* and the U.S. Centers for Disease Control and Prevention with an overall goal to strengthen the public health surveillance platform, *Sistema de Vigilancia en Salud Pública* (Sivigila). From May to November 2016, pregnant women with symptoms in three Colombian cities (Barranquilla, Cúcuta, Cali), chosen for their high rates of symptomatic Zika infection, were enrolled, and data were abstracted from pregnancy and delivery records (Table 2). Additionally, eligible women identified through Sivigila with laboratory-confirmed testing were also enrolled. Enrolled infants were followed for two years of age and evaluated for clinical outcomes.

#### Maternal Follow-Up

Upon enrollment, women provided data on their medical, pregnancy, symptom, and educational history. Any previous clinical ultrasounds and results from Zika laboratory testing during pregnancy were abstracted for the project. Throughout pregnancy, maternal serum and urine specimens were routinely collected. In the case of a live birth, umbilical cord blood and tissue, as well as placental tissue were collected. For women identified through Sivigila, a blood sample was collected as soon as possible after symptom onset. If amniotic fluid was indicated and collected for clinical reasons other than Zika, an aliquot was used for Zika testing.

As noted above, laboratory procedures for detecting Zika evolved throughout the course of the project as assays and training to perform the tests became available in Colombia. The INS aimed to collect and test maternal serum collected through Sivigila within five days of symptom onset, although this timeline was not always feasible. Depending on when the case occurred in the outbreak, samples were tested for viral RNA using one of two nucleic acid amplification tests (NAAT) procedures: an rRT-PCR singleplex assay, or the Trioplex rRT-PCR, which detects RNA from Zika, dengue, and chikungunya simultaneously. Aliquots from all samples were sent to the CDC and any remaining samples from Project VEZ were stored in a biorepository created by the INS for the Zika outbreak in Colombia. The CDC tested all maternal serum and urine samples using the Trioplex assay. Additionally, all maternal serum samples were tested for Zika IgM antibodies using the Zika virus MAC-ELISA. Samples that were positive for Zika IgM were tested for dengue IgM using the InBios DENV Detect IgM Capture ELISA. Placental, umbilical cord, and fetal tissues were tested using a conventional RT-PCR, followed by Sanger sequencing. The INS, in consultation with the CDC, used all available laboratory results (i.e., results from medical records, Sivigila, INS, and CDC) to create an algorithm to define laboratory evidence for Zika infection (Table 3).

#### Infant Follow-Up

In the case of a live birth, the infant underwent neuroimaging and an ophthalmologic examination. In the case of fetal loss, tissue was collected, and a pathology examination was performed. Zika-associated birth defects, characterized by any brain abnormalities with or without microcephaly and structural eye abnormalities, were identified within the first two weeks after birth. Microcephaly was defined as head circumference below the third percentile for age and sex. Any previous diagnoses of microcephaly in the medical records were reevaluated. Children who met these criteria only after the first two weeks postpartum were considered to have postnatal-onset microcephaly. Infants born without any Zika-associated birth defects were assessed for preterm delivery, and low birth weight. After two weeks postpartum, the presence of any neurodevelopmental outcomes, including seizures, swallowing abnormalities, tone abnormalities, movement abnormalities, arthrogryposis, as well as visual impairment and hearing abnormalities was assessed. Additionally, any potential delay in achieving developmental milestones was evaluated through the standardized Colombian screening assessment, the *Escala Abreviada de Desarrollo* (EAD-1). Results that indicated potential delay in achieving developmental milestones, also known as “alert scores,” in any of four primary developmental domains (gross motor, fine motor, personal-social, and hearing and language) were recorded. Due to outcomes such as fetal loss, follow-up data were available for 990 of the 1,225 (84%, accounting for twins) enrolled infants (Table 4).^38^

#### Key Attributes

Project VEZ enrolled a large population that included both infants who were prenatally exposed to Zika with and without Zika associated birth defects. All prenatally exposed infants had laboratory or clinical confirmation of Zika virus diagnosis. For around 40% of infants, neuroimaging, including cranial ultrasound, computed tomography, or magnetic resonance imaging, was obtained. While the research protocol included neurodevelopmental evaluations, some infants did not have follow-up.

### Zika en Embarazadas y Niños (multiple sites) – Instituto Nacional de Salud de Colombia, U.S. Centers for Disease Control and Prevention

#### Overview

The *Zika en Embarazadas y Niños* is a multi-site prospective cohort study in Colombia, established by the INS in collaboration with the CDC, aimed at assessing the risk of adverse maternal outcomes, prenatal brain injury, and postnatal neurodevelopmental abnormalities associated with Zika and other prenatal viral infections. The study enrolled 1,519 pregnant women, 287 partners, and 1,108 infants from thirteen prenatal care clinics located in three Colombian departments (Atlántico, Santander, Valle del Cauca) known for a high prevalence of Zika infection (Table 2). The enrolled pregnant women, aged 16 years or older and within the first trimester of pregnancy, were followed through pregnancy and delivery. The 1,108 enrolled infants were followed for six months postpartum and evaluated for clinical outcomes. A subset of 850 infants were followed for eighteen months for developmental evaluations.

#### Maternal Follow-Up

Upon enrollment, women provided data on their medical, pregnancy, and educational history, along with whole blood and serum samples for Zika, dengue, chikungunya, syphilis, toxoplasmosis, rubella, cytomegalovirus, and herpes I and II testing (Trioplex rRT-PCR, ZIKV IgM MAC-ELISA 1.0 and 2.0, Panbio Dengue IgM Capture ELISA, TORCH). Throughout the study, blood and urine samples were collected monthly. Symptom questionnaires were administered every two weeks. Any previous clinical ultrasounds were abstracted for the project. Maternal serum was collected upon delivery. In the case of fetal loss, placental tissue was collected.

Women experiencing at least two Zika-like symptoms were asked to provide an additional blood sample. In the case of a positive rRT-PCR Zika test, an additional blood sample was collected monthly in lieu of the urine samples until two consecutive tests were negative. Throughout the study, parents received incentives, such as developmentally appropriate toys for their child and were educated about Zika prevention, signs, symptoms, and resulting pregnancy complications (Table 3).

#### Paternal Follow-Up

Upon enrollment, men (partners of the pregnant women) provided data on their educational history, along with a blood sample. Urine samples were collected monthly. Symptom questionnaires were administered every two weeks until the start of the third trimester.

Men experiencing Zika-like symptoms were asked to provide an additional blood sample. In the case of a positive rRT-PCR Zika test, a semen sample was collected every two weeks until two consecutive tests were negative.

#### Infant Follow-Up

In the case of a live birth, venous blood samples from infants were collected for testing within the first 10 days. In the case of fetal loss, tissue was collected. Anthropometric measurements (beginning at birth) and urine samples (beginning at two weeks) were collected biweekly. Through the first six months of age, infants underwent neurological, auditory, visual, and developmental assessments. For infants enrolled in the eighteen-month follow-up, visual evaluations and developmental assessments were administered again at nine, twelve, and eighteen months of age. Three validated screening tools (EAD-3, ASQ-3, ASQ:SE-2) and one diagnostic tool (BSID-III) were used in evaluating the infant’s development (Table 4).^39^

#### Key Attributes

Project ZEN enrolled a large number of infants who were prenatally exposed to Zika and those who were not. For those who were exposed, there was laboratory and clinical confirmation of infection. Additionally, infants underwent neurodevelopmental follow-up by more than one pediatrician. However, the Zika outbreak had waned by early 2017 when enrollment in the ZEN cohort began. The Zika infection rate in this cohort was substantially lower than the incidence at the peak of the epidemic (1% for this study, compared to 10%). In anticipation of fewer affected pregnancies, clinicians conducted a review of each case enrolled in ZEN.

### Zika in Infants and Pregnancy (multiple sites) – National Institutes of Health

#### Overview

The Zika in Infants and Pregnancy (ZIP) multi-site study is a prospective, international, multisite cohort that was launched in 2016 by the National Institutes of Health to evaluate the health risks that Zika poses to pregnant women and their infants. With an initial goal of recruiting 10,000 mother-infant dyads, the study sought to compare the occurrence of prenatal brain injury and postnatal neurodevelopmental abnormalities in infected and uninfected pregnant women, as well as in symptomatic and non-symptomatic pregnant women. Pregnant women aged 15 years or older within the first or early second trimester of pregnancy in Zika-endemic regions of Brazil, Colombia, Guatemala, Nicaragua, Puerto Rico, and Peru were enrolled and followed longitudinally through pregnancy, delivery, and six weeks postpartum (Table 2). Additionally, pregnant women with symptomatic infection confirmed by serology and/or RT-PCR were also enrolled, regardless of gestational age. Women were monitored for infection through symptoms of Zika-like illness and laboratory sampling. Enrolled infants were followed through one year of age and evaluated for clinical and developmental outcomes.

#### Maternal Follow-Up

Upon enrollment, women provided data on their medical, vaccination, and pregnancy history. At each monthly visit, women had a physical examination, completed a health questionnaire regarding exposure to environmental hazards and Zika-like symptoms, and provided samples (serum, plasma, whole blood, urine, saliva, and vaginal swabs) for assessment of viral exposure and infection. In between monthly visits, women provided urine samples. Clinically indicated ultrasounds were recorded. In the case of a live birth, umbilical cord blood, placental tissue, and amniotic fluid were collected. In the case of fetal loss, placental tissue was collected.

Women experiencing Zika-like symptoms were asked to provide additional specimens for testing. In the case of positive Zika testing, two additional clinic visits were scheduled, and additional blood, urine, saliva and vaginal fluid samples were collected. Follow-up imaging using an ultrasound was also obtained. Some samples were also tested for dengue, chikungunya, West Nile Virus, toxoplasma, rubella, cytomegalovirus, syphilis, and HSV. Women were educated about Zika prevention, signs, symptoms, and pregnancy complications throughout the study (Table 3).

#### Infant Follow-Up

In the case of a live birth, infant’s peripheral blood, urine, and saliva were collected and tested for evidence of infection. In the case of fetal loss, fetal tissue was collected. Neonates underwent neurological (including auditory and visual) evaluations. After birth, infants had general physical and neurological examinations, auditory, visual, and neurodevelopmental evaluations, and laboratory testing in the first year of life. If neurological assessments proved abnormal, additional testing was performed. Any radiological data from infants with Zika exposure were recorded. If any cerebrospinal fluid was collected, excess was kept and stored in the biorepository (Table 4).^43^

#### Key Attributes

The ZIP study is a large, multi-site study that primarily enrolled non-affected participants because enrollment started as the epidemic waned, and therefore, serves as an excellent reference population. Infants were seen for neurodevelopmental follow-up.

### São Paulo - Natural History of Zika Virus Infection in Gestation – Universidade de São Paulo, Brazil

#### Overview

The Natural History of Zika Virus Infection in Gestation project is a prospective, population-based study originating in Ribeirão Preto, Brazil. Dr. Marisa M. Mussi and her team enrolled a large cohort of Zika-exposed and Zika-infected symptomatic pregnant women and their infants from the 2015-2016 outbreak. A total of 1,116 pregnant women with Zika-like symptoms gave birth in the region, and 511 were confirmed to have been infected by Zika virus. Of these, 189 were followed through pregnancy and delivery (Table 2). From this cohort, several studies were completed, as described below.

Coutinho et. al describes a study to define the prevalence of adverse outcomes after maternal Zika infection in the NATZIG cohort.^23^ The study included 511 Zika-infected women and their 513 infants (two sets of twins). A follow-up study correlated detection of anti-ZIKV-IgM and varying levels of anti-ZIKV-IgG antibodies with long-term outcomes (up to 24 months of age) among a cohort of prenatally exposed infants.^44^ The study compared 30 children with congenial Zika syndrome to 123 children who did not have adverse outcomes at birth. Teixeira et. al evaluated for signs of prenatal Zika exposure by cranial ultrasounds in 219 prenatally exposed infants from the NATZIG cohort and compared these with 170 non-exposed infants from the ZIP cohort (described above).^45^

#### Maternal Follow-Up

Pregnancy, delivery, and neonatal history and clinical evaluations were abstracted from medical records. Samples of blood and urine were collected.

Zika infection was confirmed for 511 women via ZIKV-RNA testing in samples of blood, urine, amniotic fluid, or placental tissue (Table 3).

#### Infant Follow-Up

Blood, saliva, and urine samples were collected from infants. Samples were tested for anti-IgM antibodies and for Zika RNA. Children underwent physical and neurological examinations. Birth weight and head circumference were measured and classified based on INTERGROWTH-21st criterion. At subsequent follow-up appointments, growth, neurodevelopment, and morbidity were recorded. A subset of infants born in the hospital or enrolled within six months of age underwent additional assessments, including the Hammersmith Neonatal Neurological Examination (HNNE), a pediatric evaluation, the Bayley-III, a hearing screening, neuroimaging, and an eye examination (Table 4).^23,44,45^

#### Key Attributes

The NATZIG cohort enrolled a large number of exposed and infected mother-infant dyads. Additionally, children underwent neuroimaging and neurodevelopmental follow-up.

### Rio de Janeiro - Instituto Fernandes Figueira – Oswaldo Cruz Foundation

#### Overview

*Instituto Fernandes Figueira* (IFF) in the Oswaldo Cruz Foundation (Fiocruz) is a Ministry of Health referral center for high-risk pregnancies in Rio de Janeiro, Brazil. The institution follows a large number of children with prenatal Zika exposure, which allows for several studies to be executed within the same cohort, some of which are described below. There was variation in the genetic and clinical data collected for mother-infant pairs.

Brasil et al. describe the first cohort study, done to characterize the range of outcomes that follow prenatal Zika infection. A total of 345 women were enrolled from September 2015 to May 2016 and followed through delivery. The researchers found that despite a mild clinical manifestation of infection in a mother, prenatal Zika infection can be associated with growth restriction, central nervous abnormalities, and even fetal demise.^46^

Secondly, Zin et al. describe a study done in 2016 to expand the concept of screening at that time, showing which infants exposed to Zika prenatally should be evaluated for eye abnormalities. The researchers evaluated a cohort of 112 infants and described alterations in association with Zika vertical transmission and their association with certain central nervous system findings, microcephaly, and timepoint of infection.^47^

A third paper, Cranton et. al, describes a study done to a) characterize the spectrum of outcomes associated with prenatal Zika virus and b) determine if there is an association between head circumference at birth and neurocognitive development for normocephalic infants. Based on data from the cohort of 296 infants, researchers concluded that the vast spectrum of outcomes needs to be recognized to enable early referral to treatment. Additionally, head circumference at birth was associated with neurodevelopmental outcome for children with normocephaly.^48^

#### Maternal Follow-Up

Samples collected from women include blood, urine, amniotic fluid, serum, and placental tissue.

Maternal infection was confirmed for most women. Timing of infection was defined as the week of gestation where a mother experienced Zika-like infants and/or received a positive RT-PCR result. RT-PCR was also used to test for dengue, chikungunya, CMV, and human immunodeficiency virus (HIV). Some samples were tested for antibodies to dengue and chikungunya.

#### Infant Follow-Up

Samples collected from infants include blood, cerebrospinal fluid, and urine.

Varying types of clinical data were collected from infants. Upon enrollment, demographic, medical, and prenatal history were recorded, and infants had physical exams. Anthropometric measurements (i.e., weight, length, head circumference) were taken. Some infants were evaluated with the Bayley-III. Some underwent imaging, (i.e., MRI, CT, transfontanelle US) complete eye examinations, audiology exams, and neuropsychological testing.^46-49^

#### Key Attributes

The IFF cohort is representative of the spectrum of outcomes seen after prenatal Zika infection, which allows for specificity in analysis. Additionally, children underwent neuroimaging and neurodevelopmental testing.

### Harmonizing Clinical Data

As the data collection protocols varied in each study, creating a centralized database that both compartmentalizes each site’s information and integrates data across the entire study became crucial for data analysis.^50^ To harmonize the data, we designed an iterative four-step process to integrate clinical, laboratory, and radiological data into a single database where information is harmonized and interpretable for the cohort as a whole (Figure 3). We began by reviewing the data obtained from each site to identify overlapping variables. Received information was then categorized as an exposure, modifier, or outcome, and converted into categorical criteria. For example, an outcome relating to microcephaly was entered as “present” or “absent.” When available, we also kept quantitative data, such as head measurements, as continuous variables. We housed the data in a secure Research Electronic Data Capture (REDCap) database. We evaluated data transfer for functionality and reliability, and all entered data was cross-reviewed and validated by at least two members of our research team.

**Figure 3.**
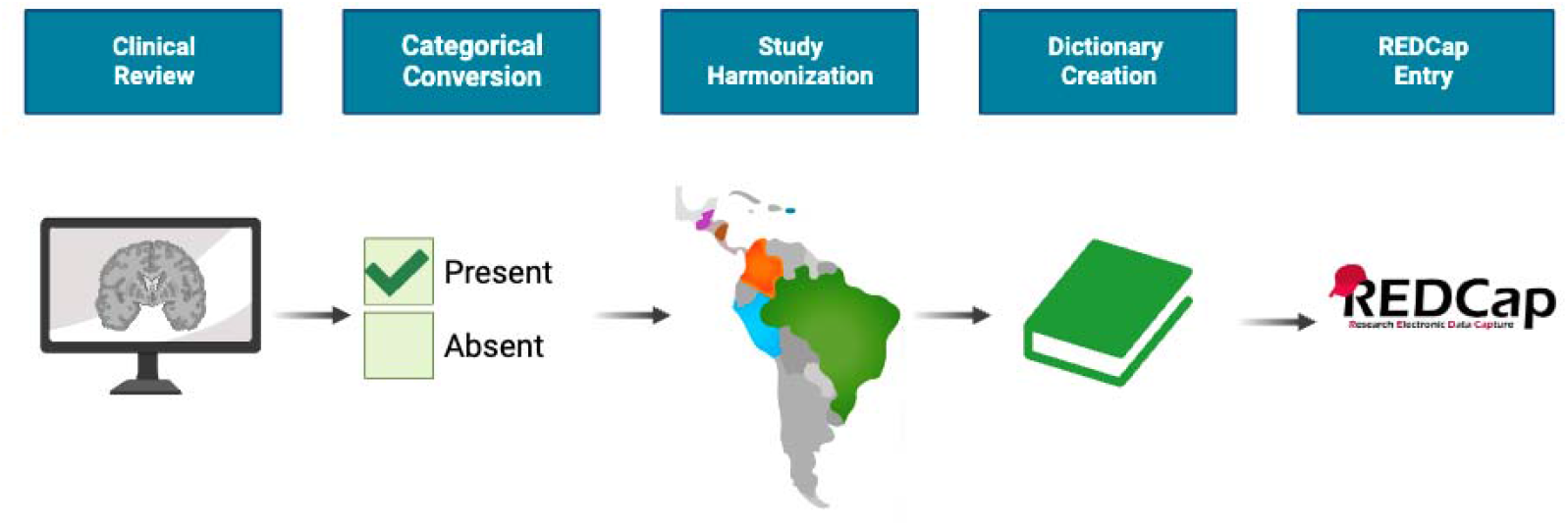
Harmonizing Study Data. Clinical data from each study was reviewed and converted into a categorical format. Then, data from each study site was aligned across the cohorts. Next, a data dictionary was created to provide a key for importing the clinical outcomes into the database. Lastly, participant information was imported into REDCap to evaluate its operational capabilities. Finally, the data was imported in a de-identified manner and validated.

The four domains of collected data include 1) demographics; 2) clinical evaluations; 3) laboratory testing; and 4) radiological findings (Figure 4). Among these, clinical outcome and laboratory testing information were among the most detailed and variable. We prioritized data extraction and management for such information. Radiological information included study type, age of imaging, and bodily system. Criteria for categorization of imaging findings was reviewed by a neuroradiologist.

**Figure 4.**
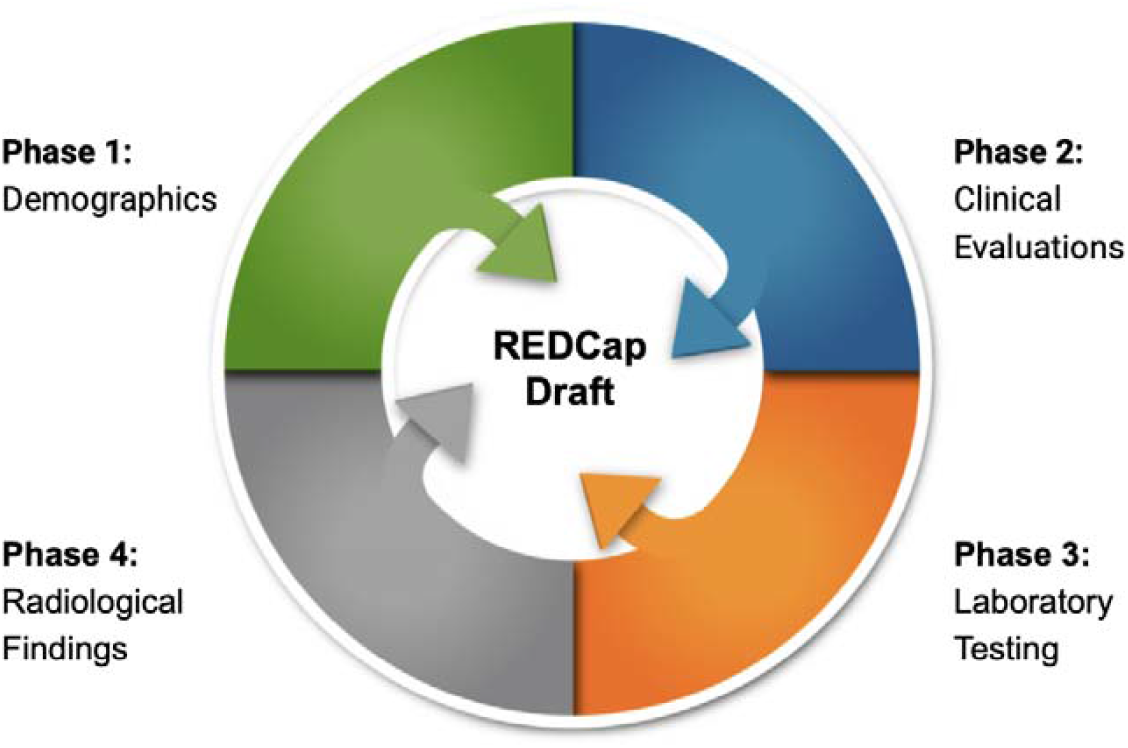
REDCap Creation Plan. Participant’s demographics, clinical evaluations, laboratory testing, and radiological findings were reviewed among the studies and iteratively aligned per the Study Data Harmonization process outlined in Figure 3. As radiological findings and clinical evaluations contained the most variable information across studies, these phases were prioritized during REDCap drafting.

## EARLY ASSESSMENT OF THE ADOPTED RESEARCH STRATEGY

The PING Consortium’s current study has key strengths and limitations that we addressed, as described below.

### Establishing a Repository

The Consortium brings together existing cohorts evaluated with clinical phenotyping and pre- and postnatal neuroimaging. In addition, multiple biological specimens were collected. Together, these resources enable genomic association studies and are a resource for the scientific community in years to come. In addition to Zika-related information, partnering cohorts also provided clinical and laboratory data evaluating for other prenatal viral infections, including CMV. As such, the repository is critically important in facilitating future research projects and can be centrally accessible for investigators. Protocols for accessing the data and samples are being developed by the Consortium, as permitted by parent studies.

### Bringing Together Different Types of Studies

There are several advantages in bringing together participants enrolled in single- and multi-site research programs in different countries. First, our study benefits from access to multiple, diverse, and traditionally underserved and understudied populations, from urban to rural, and in various countries. Larger and more diverse participant cohorts add to the generalizability and durability of research findings. Second, the inclusion of prospective and retrospective data provides a wealth of cross-sectional and longitudinal data, from timepoints before and after the epidemics. Finally, a larger cohort provided more statistical power in identifying genetic associations for common and rare variants.

### Protecting Participants While Adding Value to Research Conducted

Parent studies were approved at local and/or national IRBs, and therefore utilized appropriate measures to safeguard the rights of the participant. Such safeguards included obtaining informed consent and deidentifying collected data. A wealth of data collected in parent studies is directly applicable to the described research program, at minimal additional risk to the participants. The Consortium also implemented strict data management procedures that ensure data integrity and participant protection while facilitating data analysis and dissemination of research findings. Combined, these measures preserve the anonymity and safety of participants while establishing an invaluable platform for collaboration and progress in the field, all with previously collected, available, and otherwise utilized and completed human studies.

### Study Limitations

In addition to the logistical complexities of coordinating a multi-site research program, there were limitations that needed to be addressed. These are described below.

### Differences Among Studies

Our goal is to investigate if genetic variants are associated with virally induced prenatal brain injury in diverse participant cohorts. However, there was heterogeneity in the clinical data collected among different cohort studies. When similar datasets were collected, there were variations in data collection protocols. The variability is addressed, in part, by harmonizing the datasets, as described above. However, there were unique and site-specific interests that did not always align, and these datasets were integrated when possible. For instance, while the focus of radiologists’ impressions in the Puerto Rico study focused on craniofacial abnormalities, access to the CT and MRI images enables us to assess the whole brain and extract new insights.

### Availability of Biospecimen

Studies had varying sample collection and storage protocols. As a result, samples obtained were not always optimized for DNA extraction and genome sequencing. In some cases, differences in sample storage or processing conditions contributed to DNA degradation. As a result, only a subset of the cohort can be included in the genetic association study.

### IRB Review

Institutional Review Board approval was time costly because protocols were evaluated by each site separately and the requirements for approval did not always align. In several cases, regional or national review and approval was additionally required. Such review added time in performing the research and was a resource-intensive process.

### Logistics of Transporting Samples

The large-scale, international nature of this collaboration introduced logistical considerations, especially with respect to coordinating sample transport. Despite efforts to standardize sample transport protocols, variations in transit conditions and transport times across sites were unavoidable. We obtained a CDC import permit and set up a service agreement with courier services to support partners in transporting samples.

## Data Availability

All data produced in the present study are available upon reasonable request to the authors.

## SUMMARY AND FUTURE WORK

Establishing the Consortium as a network of international investigators and sites is a collaborative effort, laying the foundation for multi-center research initiatives to address fundamental, unanswered questions in the fields of maternal and fetal health, prenatal brain development and risk for viral infections. Here, we describe the formation of the Prenatal Infection and Neurodevelopmental Genetics (PING) Consortium and the present study design, developed to advance a conceptual understanding of virally induced prenatal brain injury.

The Consortium is continuing to establish partnerships with sites with active studies to further expand the repository of maternal and fetal clinical data and biospecimen. Further, select member sites located in endemic regions are actively recruiting participants and/or completing follow-up studies. Ongoing work also includes performing genomic association analyses to identify maternal and fetal modifiers in developing prenatal brain injury.

### LIST OF ABBREVIATIONS

PING: Prenatal Infection and Neurodevelopmental Genetics Consortium

## DECLARATIONS

### Ethics Approval and Consent to Participate

Samples and associated data collected through the Prenatal Infection and Neurodevelopmental Genetics Consortium were IRB approved by participating institutions and by Children’s National Hospital (IRB reference number 8259). Any changes to the protocol or materials are submitted for approval by the IRB/CEP before being implemented, through amendments to the project. The research team notifies the IRB/CEP of deviations from the protocol or any adverse events that might be related to the present study. Brazilian studies were also approved by CONEP (approval numbers of the original related projects: CAAEs: 61936216.9.0000.5404; 56673616.3.2002.5440; 61936216.9.0000.5404).

The researchers ensured that this study was conducted in full compliance with the principles set out in the Belmont Report: Ethical Principles and Guidelines for the Protection of Human Subjects in Research by the US National Commission for the Protection of Human Subjects in Biomedical and Behavioral Research (18 April 1979) and encoded in 45 CFR Section 46 or the ICH E6; 62 Federal Regulations 25691 (1997). The Investigators’ institution must maintain an up-to-date, federal-level policy (FWA) issued by the Office of Human Protection in Research for US government-funded research.

Both multi-center and site-specific IRB approvals were obtained in one of in two ways. A majority of the studies obtained informed consent for genetic testing prospectively. In some cases, informed consent was obtained retrospectively.

### Consent for Publication Not applicable

Availability of Data and Materials Not applicable.

## Authors’ Contributions

YK, CA, SS, and BF wrote the first draft of the manuscript. All co-authors reviewed and edited the manuscript, and edits were reviewed and incorporated by CA, BF, and YK. YK supervised the work. Co-authors reviewed and approved the final version of the manuscript.

## Acknowledgements

We thank research participants and their families, and the faculty and staff at our institutions. We are also grateful for the support of Children’s National Hospital for translational research.

## Supplementary Information

**Supplemental Figure 1.**
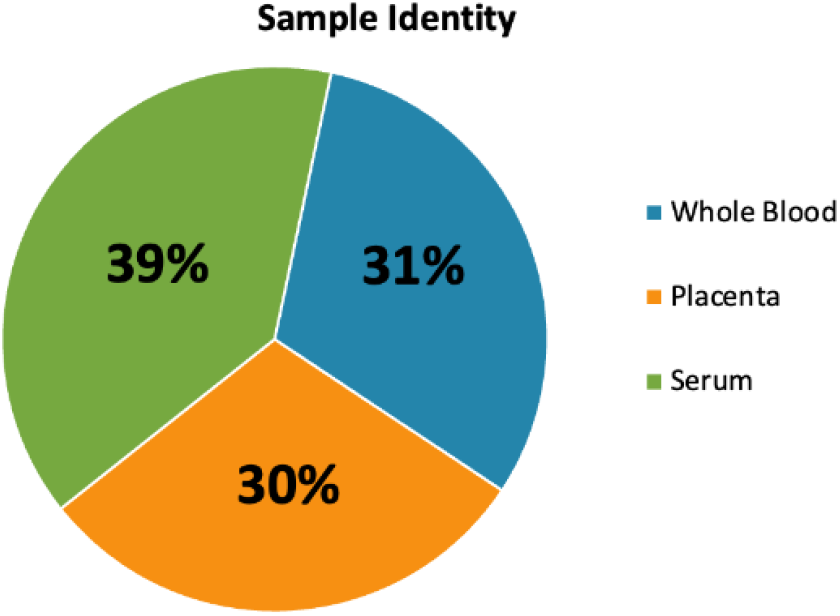
Sample Identity of Biological Samples. The biorepository is comprised of a variety of sample identities. This includes blood (31%), placental tissue (30%) and serum (39%). All samples are undergoing DNA extraction for sequencing, analysis, and storage.

**Supplemental Table 1.**
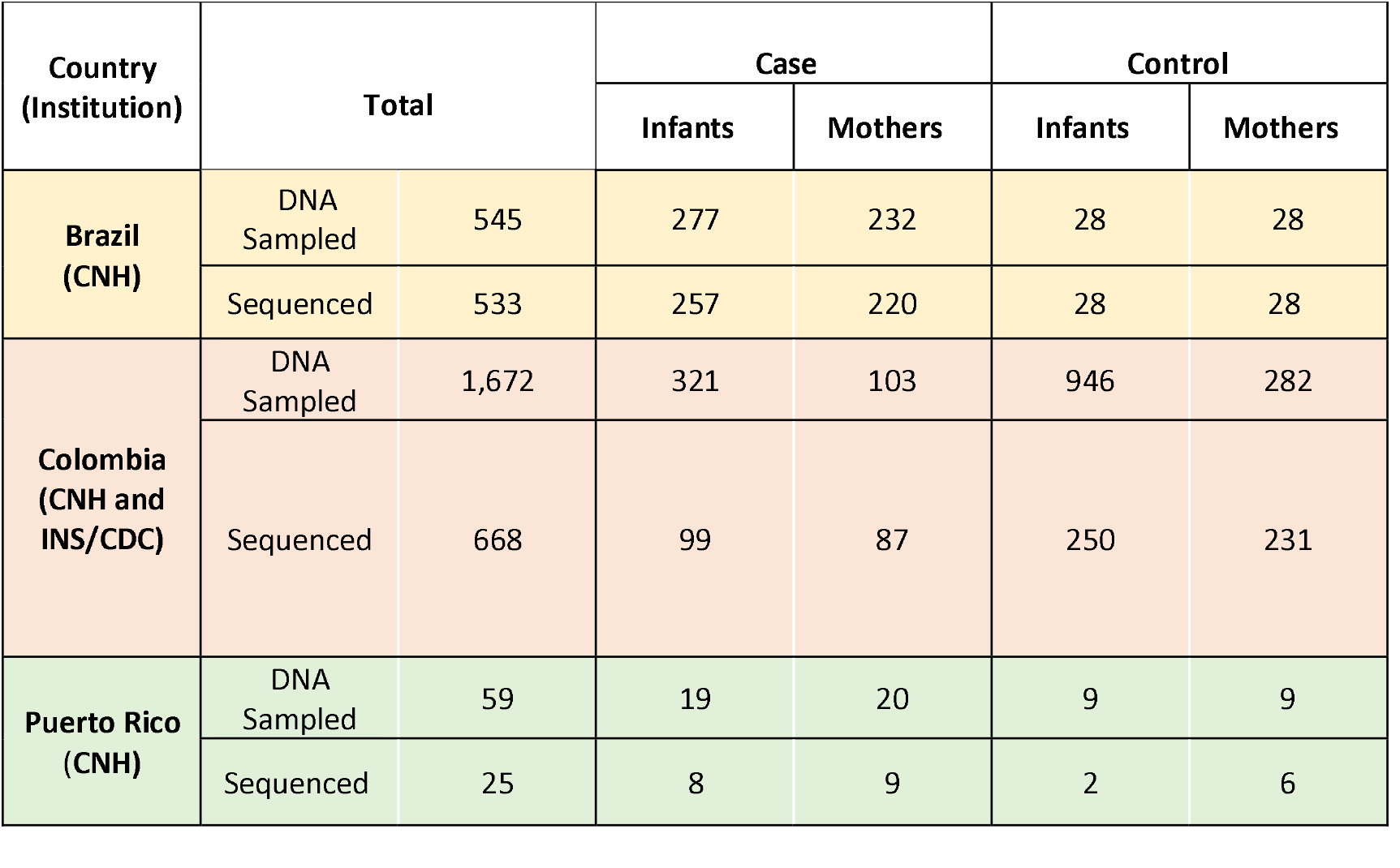
Status of Sequencing for Samples Currently in the Consortium’s Biorepository.

| Country<br>(Institution) | Total |  | Case |  | Control |  |
| --- | --- | --- | --- | --- | --- | --- |
|  |  |  | Infants | Mothers | Infants | Mothers |
| Brazil<br>(CNH) | DNA<br>Sampled | 545 | 277 | 232 | 28 | 28 |
|  | Sequenced | 533 | 257 | 220 | 28 | 28 |
| Colombia<br>(CNH and<br>INS/CDC) | DNA<br>Sampled | 1,672 | 321 | 103 | 946 | 282 |
|  | Sequenced | 668 | 99 | 87 | 250 | 231 |
| Puerto Rico<br>(CNH) | DNA<br>Sampled | 59 | 19 | 20 | 9 | 9 |
|  | Sequenced | 25 | 8 | 9 | 2 | 6 |

## BIBLIOGRAPHY

1. Rudbeck Roge, H. & Henriques, U. Fetal and perinatal infections. A consecutive study. Pathol Res Pract 188, 135–40 (1992).

2. Khan, A.M., Morris, S.K. & Bhutta, Z.A. Neonatal and Perinatal Infections. Pediatr Clin North Am 64, 785–798 (8017).

3. Lynch, L. & Ghidini, A. Perinatal infections. Curr Opin Obstet Gynecol 5, 24–32 (1993).

4. Edwards, A.D. & Tan, S. Perinatal infections, prematurity and brain injury. Curr Opin Pediatr 18, 119–24 (2006).

5. Velu, P.P. et al. Epidemiology and aetiology of maternal bacterial and viral infections in low- and middle-income countries. J Glob Health 1, 171–88 (2011).

6. Racicot, K. & Mor, G. Risks associated with viral infections during pregnancy. J Clin Invest 127, 1591–1599 (2017).

7. Mulkey, S.B. et al. Neurodevelopmental Abnormalities in Children With In Utero Zika Virus Exposure Without Congenital Zika Syndrome. JAMA Pediatr 174, 269–276 (2020).

8. Counotte, M.J. et al. Zika virus infection as a cause of congenital brain abnormalities and Guillain-Barre syndrome: A living systematic review. F1000Res 8, 1433 (2019).

9. Kimberlin, D.W. et al. Valganciclovir for symptomatic congenital cytomegalovirus disease. N Engl J Med 372, 933–43 (2015).

10. Kousa, Y.A. & Hossain, R.A. Causes of Phenotypic Variability and Disabilities after Prenatal Viral Infections. Trop Med Infect Dis 6 (2021).

11. Counotte, M.J. et al. Zika virus infection as a cause of congenital brain abnormalities and Guillain-Barre syndrome: From systematic review to living systematic review. F1000Res 7, 196 (2018).

12. Kundakovic, M. & Jaric, I. The Epigenetic Link between Prenatal Adverse Environments and Neurodevelopmental Disorders. Genes (Basel) 8 (2017).

13. Fellay, J. et al. A whole-genome association study of major determinants for host control of HIV-1. Science 317, 944–7 (2007).

14. Fellay, J. et al. Common genetic variation and the control of HIV-1 in humans. PLoS Genet 5, e1000791 (2009).

15. Trachtenberg, E. et al. The HLA-B/-C haplotype block contains major determinants for host control of HIV. Genes Immun 10, 673–7 (2009).

16. Shan, C. et al. A Zika virus envelope mutation preceding the 2015 epidemic enhances virulence and fitness for transmission. Proc Natl Acad Sci U S A 117, 20190–20197 (2020).

17. Moore, C.A. et al. Characterizing the Pattern of Anomalies in Congenital Zika Syndrome for Pediatric Clinicians. JAMA Pediatr 171, 288–295 (2017).

18. Haig, D. Genetic conflicts in human pregnancy. Q Rev Biol 68, 495–532 (1993).

19. Paixao, E.S., Leong, W.Y., Rodrigues, L.C. & Wilder-Smith, A. Asymptomatic Prenatal Zika Virus Infection and Congenital Zika Syndrome. Open Forum Infect Dis 5, ofy073 (2018).

20. Santos, C.N.O. et al. Association Between Zika Virus Microcephaly in Newborns With the rs3775291 Variant in Toll-Like Receptor 3 and rs1799964 Variant at Tumor Necrosis Factor-alpha Gene. J Infect Dis 220, 1797–1801 (2019).

21. Mercado, M. et al. Discordant Clinical Outcomes in a Monozygotic Dichorionic-Diamniotic Twin Pregnancy with Probable Zika Virus Exposure. Case Report. Trop Med Infect Dis 5 (2020).

22. Caires-Junior, L.C. et al. Discordant congenital Zika syndrome twins show differential in vitro viral susceptibility of neural progenitor cells. Nat Commun 9, 475 (2018).

23. Coutinho, C.M. et al. Early maternal Zika infection predicts severe neonatal neurological damage: results from the prospective Natural History of Zika Virus Infection in Gestation cohort study. BJOG 128, 317–326 (2021).

24. Chavali, P.L. et al. Neurodevelopmental protein Musashi-1 interacts with the Zika genome and promotes viral replication. Science 357, 83–88 (2017).

25. Han, D. et al. Human Cytomegalovirus IE2 Protein Disturbs Brain Development by the Dysregulation of Neural Stem Cell Maintenance and the Polarization of Migrating Neurons. J Virol 91 (2017).

26. Borda, V. et al. Whole-exome sequencing reveals insights into genetic susceptibility to Congenital Zika Syndrome. PLoS Negl Trop Dis 15, e0009507 (2021).

27. Aguiar, R.S. et al. Molecular alterations in the extracellular matrix in the brains of newborns with congenital Zika syndrome. Sci Signal 13 (2020).

28. Linden, V.V. et al. Discordant clinical outcomes of congenital Zika virus infection in twin pregnancies. Arq Neuropsiquiatr 75, 381–386 (2017).

29. Santos, V.S. et al. Case Report: Microcephaly in Twins due to the Zika Virus. Am J Trop Med Hyg 97, 151–154 (2017).

30. Sobhani, N.C. et al. Discordant Zika Virus Findings in Twin Pregnancies Complicated by Antenatal Zika Virus Exposure: A Prospective Cohort. J Infect Dis 221, 1838–1845 (2020).

31. Bulik-Sullivan, B.K. et al. LD Score regression distinguishes confounding from polygenicity in genome-wide association studies. Nat Genet 47, 291–5 (2015).

32. Tanaka, T. et al. Genome-wide meta-analysis of observational studies shows common genetic variants associated with macronutrient intake. Am J Clin Nutr 97, 1395–402 (2013).

33. Levy, D. et al. Genome-wide association study of blood pressure and hypertension. Nat Genet 41, 677–87 (2009).

34. Zimon, M. et al. Pairwise effects between lipid GWAS genes modulate lipid plasma levels and cellular uptake. Nat Commun 12, 6411 (2021).

35. Neu, N., Duchon, J. & Zachariah, P. TORCH infections. Clin Perinatol 42, 77–103, viii (2015).

36. Wolford, R.W. & Schaefer, T.J. Zika Virus. in StatPearls (Treasure Island (FL), 2023).

37. Pielnaa, P. et al. Zika virus-spread, epidemiology, genome, transmission cycle, clinical manifestation, associated challenges, vaccine and antiviral drug development. Virology 543, 34–42 (2020).

38. Mercado-Reyes, M. et al. Pregnancy, Birth, Infant, and Early Childhood Neurodevelopmental Outcomes among a Cohort of Women with Symptoms of Zika Virus Disease during Pregnancy in Three Surveillance Sites, Project Vigilancia de Embarazadas con Zika (VEZ), Colombia, 2016-2018. Trop Med Infect Dis 6 (2021).

39. Gonzalez, M. et al. Cohort profile: congenital Zika virus infection and child neurodevelopmental outcomes in the ZEN cohort study in Colombia. Epidemiol Health 42, e2020060 (2020).

40. Van der Auwera, G.A., O’Connor, B.D. & Safari, a.O.R.M.C. Genomics in the cloud : using Docker, GATK, and WDL in Terra, (O’Reilly Media, Sebastopol, CA, 2020).

41. Mulkey, S.B. et al. Preschool neurodevelopment in Zika virus-exposed children without congenital Zika syndrome. Pediatr Res 94, 178–184 (2023).

42. de Oliveira, D.N. et al. Inflammation markers in the saliva of infants born from Zika-infected mothers: exploring potential mechanisms of microcephaly during fetal development. Sci Rep 9, 13606 (2019).

43. Lebov, J.F. et al. International prospective observational cohort study of Zika in infants and pregnancy (ZIP study): study protocol. BMC Pregnancy Childbirth 19, 282 (2019).

44. Coutinho, C.M. et al. Persistence of Anti-ZIKV-IgG over Time Is Not a Useful Congenital Infection Marker in Infants Born to ZIKV-Infected Mothers: The NATZIG Cohort. Viruses 13 (2021).

45. Reis Teixeira, S. et al. Cranial US in Infants Exposed to Zika Virus: The NATZIG Cohort. Radiology 300, 690–698 (2021).

46. Brasil, P. et al. Zika Virus Infection in Pregnant Women in Rio de Janeiro. N Engl J Med 375, 2321–2334 (2016).

47. Zin, A.A. et al. Screening Criteria for Ophthalmic Manifestations of Congenital Zika Virus Infection. JAMA Pediatr 171, 847–854 (2017).

48. Cranston, J.S. et al. Association Between Antenatal Exposure to Zika Virus and Anatomical and Neurodevelopmental Abnormalities in Children. JAMA Netw Open 3, e209303 (2020).

49. Lopes Moreira, M.E. et al. Neurodevelopment in Infants Exposed to Zika Virus In Utero. N Engl J Med 379, 2377–2379 (2018).

50. Carabali, M., Maxwell, L., Levis, B., Shreedhar, P. & Consortium, Z.I.-M. Heterogeneity of Zika virus exposure and outcome ascertainment across cohorts of pregnant women, their infants and their children: a metadata survey. BMJ Open 12, e064362 (2022).

